# Brain network hypersensitivity underlies pain crises in sickle cell disease

**DOI:** 10.1101/2023.10.08.23296715

**Authors:** Pangyu Joo, Minkyung Kim, Brianna Kish, Vidhya Vijayakrishnan Nair, Yunjie Tong, Steven E Harte, Richard E Harris, UnCheol Lee, Ying Wang

## Abstract

Sickle cell disease (SCD) is a genetic disorder causing blood vessel blockages and painful Vaso-occlusive crises (VOCs). VOCs, characterized by severe pain due to blocked blood flow, are recurrent and unpredictable, posing challenges for preventive strategies. In this study we propose explosive synchronization (ES), a phenomenon characterized by abrupt brain network phase transitions, as a novel approach to address this challenge. We hypothesized that the accumulated disruptions in the brain network induced by SCD might lead to strengthened ES and hypersensitivity. We explored ES’s relationship with patient reported outcome measures (PROMs) and VOCs by analyzing EEG data from 25 SCD patients and 18 matched controls. SCD patients exhibited significantly lower alpha wave frequency than controls. SCD patients under painful pressure stimulation showed correlation between frequency disassortativity (FDA), an ES condition, and three important PROMs. Furthermore, patients who had a higher frequency of VOCs in the preceding 12 months presented with stronger FDA. The timing of VOC occurrence relative to EEG recordings was significantly associated to FDA. We also conducted computational modeling on SCD brain network to study FDA’s role in network sensitivity. Stronger FDA correlated with higher responsivity and complexity in our model. Simulation under noisy environment showed that higher FDA could be linked to increased occurrence frequency of crisis. This study establishes connections between SCD pain and the universal network mechanism, ES, offering a strong theoretical foundation. This understanding will aid predicting VOCs and refining pain management for SCD patients.

## 1. Introduction

Sickle cell disease (SCD) is a common inherited blood disorder afflicting 1 in 500 Black Americans. Pain in SCD is a lifelong major complication starting from infancy, and can be acute, chronic, or a mixture of both. Vaso-occlusive crises (VOCs) associated with SCD are extremely painful episodes that are recurrent, unpredictable, and frequently require hospitalization and opioids for pain control. Individuals with SCD that require high doses of opioids for VOCs have an increased risk of overdose and death, development of opioid-induced hyperalgesia, and lowered quality of life (QoL) [22]. The frequency of VOCs varies widely across SCD individuals with a recent systematic review reporting a range of 0 to 18 per year [48]. Recent studies suggest that the occurrence of VOCs is associated with abnormal vasoconstriction patterns resulting from dysfunction of the autonomic nervous system, endothelial dysfunction, and/or psychological stress [8,40,43]. Central nervous system (CNS) factors may also contribute to VOCs. For example, we showed that brain connectivity between the Default Mode Network (DMN) and structures of the Salience network were altered in SCD patients that had more VOCs [47]. Interestingly, this pattern of connectivity was consistent with that observed in fibromyalgia, a chronic widespread pain disorder accompanied by fatigue, sleep disturbance, and sensory hypersensitivity [19,20].

Explosive synchronization (ES) is a universal physical phenomenon wherein a small perturbation to a network can lead to an abrupt state transition. The typical properties of an ES network include its high sensitivity to external stimuli and abrupt state transition at a tipping point [18,32]. Recent studies have suggested a link between ES in brain networks and chronic pain. We recently demonstrated that patients with fibromyalgia exhibit characteristics of ES on electroencephalography (EEG) that are associated with increased pain intensity [32], and that modulation of brain networks can transform a sensitive network into an insensitive one, or one not prone to ES [27]. These modeling investigations provided valuable insights into the network-level mechanisms of fibromyalgia and provide a rationale for studying ES mechanisms in other pain populations such as SCD.

In this study, we sought to extend our exploration of ES mechanisms into SCD. We performed EEG on SCD patients and matched healthy controls. We examined differences in ES strength in SCD patients at various times before and after VOC events. Additionally, we developed a computational model to examine the relationship between ES and the occurrence of VOCs in the SCD brain.

## 2. Methods

### 2.1 Study design and participants

Thirty-two patients with SCD (16 male and 16 female) aged 14 to 73 years, and 18 pain-free ethnicity-, age– and gender-matched healthy controls (HCs) without SCD, were enrolled. Detailed inclusion and exclusion criteria can be found in Supplementary Table 1. In brief, the main inclusion criteria for SCD participants included: 1) experiencing chronic pain greater than or equal to 3/10 on most days for the past 6 months or at least one VOC in the past 12 months, 2) no recent history of initiating or adjusting the dose of stimulant medications in the past one month, and 3) willingness to maintain current SCD and pain treatments, and not to introduce any treatment for pain for the duration of study participation.

Each participant underwent an EEG recording following the administration of patient reported outcome measures (PROMs) and routine laboratory tests for complete blood cell count, reticulocytes, and hemoglobin electrophoresis. We were unable to collect quality EEG data from some SCD patients with thick curly hair that interfered with cap placement or those experiencing a pain flare on the day of recording, leading to increased noise during EEG measurements. EEG signal noise was primarily addressed during preprocessing; however, some recordings (n=7) with severe noise could not be corrected and were excluded from analysis. Therefore, the final analysis sample was 25 (See Supplementary Figure 1). The study was approved by the Institutional Review Board at Indiana University. All patients provided written informed consent before the study.

### 2.2 Concurrent EEG with pressure pain stimulator

EEG was performed in a quiet room using multi-channel EEG equipment (Brain Products, Gilching, Germany) and a noninvasive cutaneous electrode cap with conductive gel. Recordings were done with eyes-closed during a state of rest or during concurrent pressure pain stimulation (individually calibrated to evoke moderate pain defined as a pain rating of ∼40/100, i.e., Pain40) that was elicited by a computerized pressure cuff (Hokanson, Bellevue, WA, USA) attached to the left gastrocnemius muscle. This method has been used previously to elicit evoked pressure pain in fibromyalgia patients [26]. First a resting state EEG was collected while participants rested with their eyes closed for 5 min. EEG was then recorded for 5-min during tonic pressure pain with eyes closed. Following the pressure pain protocol, resting state EEG recording was repeated for 5-min with eyes closed.

### 2.3 Patient reported outcome measures (PROMs)

Pain (pain intensity and interference) and physical function were assessed before EEG recordings with the Brief Pain Inventory (BPI) [6] and PROMIS-29 [10]. The Fibromyalgia Survey Questionnaire (FSQ), which consists of the Widespread Pain Index and the Symptom Severity scale [9,14], was utilized as a surrogate measure of nociplastic pain. Anxiety and depression were evaluated using Hospital Anxiety and Depression Scale (HADS) [6]. The number of patient-reported VOCs in the preceding 12 months was documented, a method which is commonly used in pain research for SCD [11,47]. Pain-related QoL was evaluated using the Pediatric Quality of Life Inventory (PedsQL) [35].

### 2.4 EEG analysis

#### 2.4.1 EEG preprocessing

In this study, we analyzed a 31 channel EEG (FP1, FP2, F3, F4, C3, C4, P3, P4, O1, O2, F7, F8, T7, T8, P7, P8, FZ, CZ, PZ, OZ, FC1, FC2, CP1, CP2, FC5, FC6, CP5, CP6, TP9, TP10 and POz), excluding an ECG channel (Fig.1A). The raw EEG data were preprocessed using MATLAB and the EEGLAB toolbox [13]. First, a 0.5-59Hz bandpass filter was applied to remove frequency bands with high levels of noise. Next, a visual inspection of the waveform and spectrogram of EEG was performed to remove channels with obvious noise and to segment the data into 240-second intervals. To minimize errors in spherical spline interpolation, data with more than 20% of channels removed were excluded in this study. Also, EEG data with a duration of less than 240 seconds was excluded from the analysis. With these criteria, data from 7 individuals were excluded out of the 32 SCD subjects, leaving 25 subjects’ data for analysis. Among the HC group, no data was excluded with these criteria, leaving 18 subjects’ data for analysis. Next, Independent components with a significantly high probability (>95%) being non-EEG were removed using runica in EEGLAB and IClabel [36]. The removed channels were recovered using spherical spline interpolation and finally the EEG was average referenced. In this study, we focused on analyzing alpha rhythms, which display distinct oscillatory patterns around 10Hz and are known to be highly relevant to cognitive phenomena, including pain [17,30]. To achieve this, we utilized a band-pass filter and specifically analyzed the alpha band containing 7-13Hz EEG signals.

**Figure 1.**
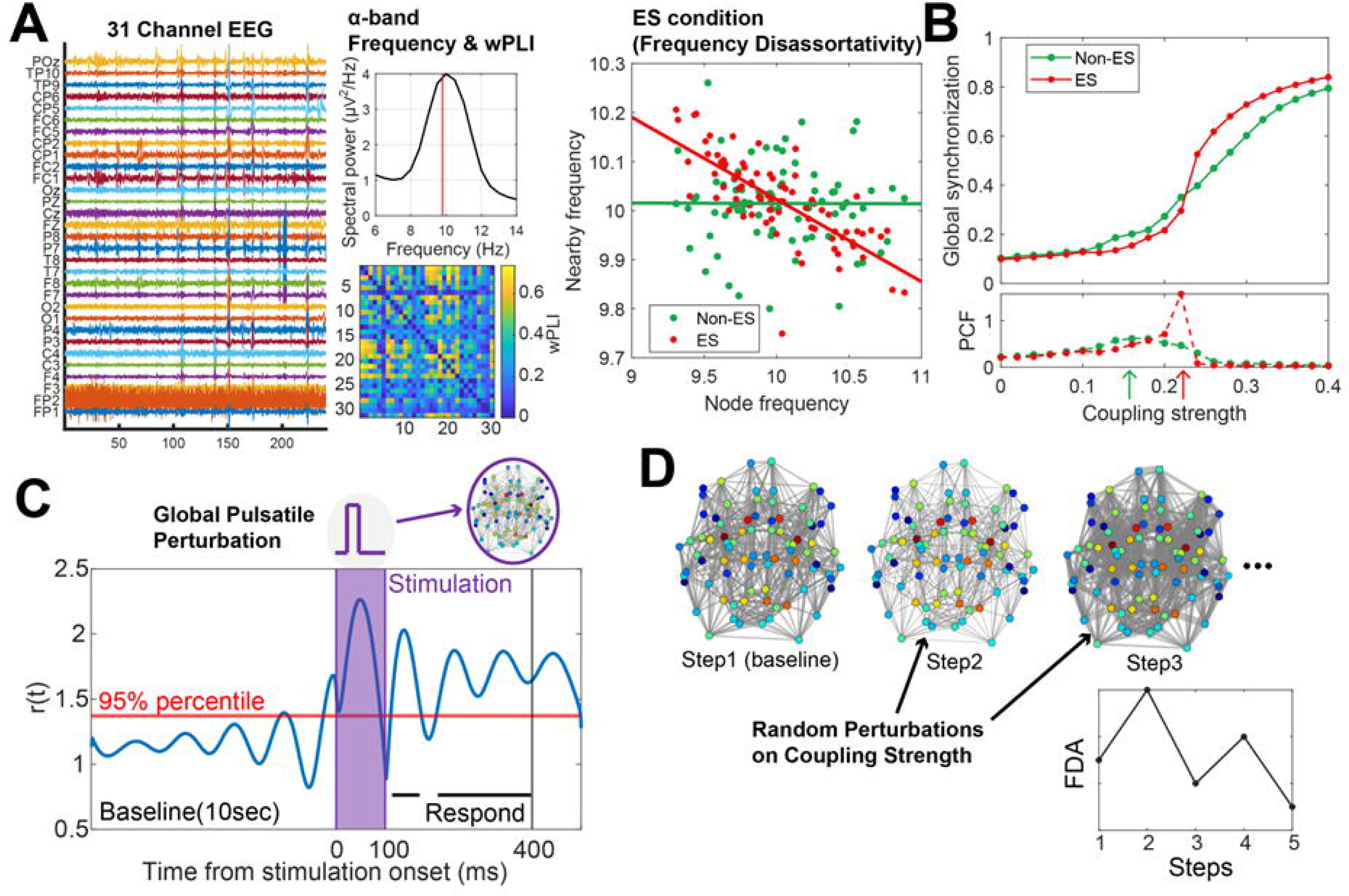
Schematic diagram of the study. A) Schematic for the EEG analysis in this study. wPLI matrix and median frequency of alpha band were calculated from 31 channel EEG. FDA, one of the ES conditions, was obtained from wPLI network and median alpha frequency. B) An example of synchronization characteristics of ES and non-ES network. The coupling strength at which the PCF reaches its maximum value was defined as critical point (green and red arrows). C) An anatomical brain network and Stuart-Landau model were used to investigate the sensitivity and frequency of state transition of the brain network. Global Pulsatile perturbation is given for 100ms and the respond from the perturbation is measured from the subsequent 300ms. D) Random perturbation on coupling strength is given to simulate number of state transitions under noisy environment.

#### 2.4.2 Weighted phase lag index (wPLI)

Weighted phase lag index (wPLI) is used to assess functional connectivity from the phase difference between two time series and is particularly resistant to volume conditions commonly observed in EEG [45]. The wPLI between the ith and jth channels is defined by the following equation.

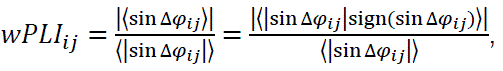

where Δφ_*ij*_ represents phase difference between ith and jth channel band EEG. The wPLI of alpha band (7-13Hz) EEG was computed with 30 sec windows and then averaged across 300 sec.

#### 2.4.3 Frequency disassortativity (FDA)

Frequency disassortativity (FDA) is a well-known ES condition that refers to the tendency where high-frequency nodes are connected to low-frequency nodes, or vice versa, within a network [34,49]. Networks that exhibit FDA have significant differences in oscillation frequencies between adjacent nodes, which can disturb global synchronization. The suppressed synchronization due to FDA can suddenly rise (Fig. 1B), a phenomenon known as ES, with an increase in network coupling strength or external perturbations. In order to measure FDA from EEG, we employed the median frequencies derived from the alpha bands and the binarized wPLI network, considering only the highest 30% wPLI in the wPLI network connectivity. Using the binarized wPLI network, the average of the adjacent median alpha frequencies for each node can be defined. Next, since we have the median alpha frequency and the average of the adjacent median alpha frequencies for each node, we can calculate the Spearman correlation between the two frequencies (ρ_*f*_). Finally, we define FDA as the negative ρ_*f*_, which signifies anti-correlation between the frequencies of connected nodes.

Defining a representative frequency for each EEG channel is important for the calculation of FDA. However, defining a representative frequency of the alpha frequency band (7-13Hz) with a peak value in alpha power spectrum may not provide an accurate result in cases where a clear peak is absent or when there is strong low-frequency activity in the power spectrum of the alpha frequency band. Therefore, in this study, median frequency was used to define each channel’s alpha frequency. The spectral power of EEG signal is calculated using Welch’s method with 2-sec window and 1-sec overlaps and the median alpha frequency is defined as the 50% percentile of cumulative spectral power within the alpha band (7-13Hz).

### 2.5 Computational modeling of SCD brain network

#### 2.5.1 Stuart-Landau based brain model

We performed a computational model of the SCD brain in order to simulate altered ES function and test for sensitivity of network mechanisms. Stuart-Landau model is a mathematical model that describes the dynamics of coupled oscillator systems and their synchronization. The Stuart-Landau oscillator is extensively utilized for simulating the oscillatory dynamics between neural masses in the brain and for simulating brain signals from many different modalities [3,12,27]. In this study, we aim to investigate the sensitivity changes in the brain and the occurrence of crises using the Stuart-Landau model.

The coupled Stuart-Landau model is defined by the following equations.

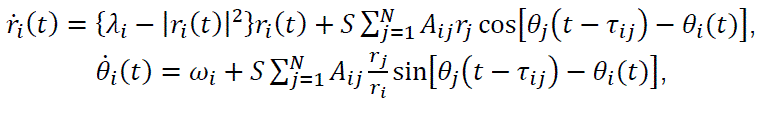

where r_*i*_(*t*) and θ_*i*_(*t*) are the amplitude and phase of ith oscillator, respectively. λ_*i*_ is a parameter that determines the size of the limit cycle of the oscillator, and it was set to 1. ω_*i*_ determines the natural frequency of an ith oscillator and was normally distributed with 10Hz mean and 0.4Hz standard deviation to simulate alpha waves in the brain. *A*_*ij*_ represents the anatomical connectivity derived from diffusion tensor imaging (DTI), which has been parcellated into 82 nodes [21]. *A*_*ij*_ was binarized, limiting its values to either 0 or 1. The time delay τ_*ij*_ = *D*_*ij*_/υ represents the delay between region j and k, where *D*_*jk*_ denotes the distance between ith and jth brain regions. Here, υ is set to 7 ms, representing the average speed of axons across brain regions [5]. *S* represents the coupling strength between oscillators and S varied from 0 to 1.

Based on the hypothesis that brain networks are in a critical state for efficient information processing [1,24,28,44], we assume that the brain network in a conscious state resides at a critical point. Also, an ES network exhibits a sharp and strong PCF peak (Fig. 1B), which can result in high sensitivity to external perturbations near the critical point. Accordingly, we adjusted the coupling strength *S* to ensure that the system is at the critical point. The patterns of synchronization and the strength of ES vary significantly depending on how the initial frequencies are distributed in a brain network. By varying *S* in increments of 0.005, the critical point that maximizes the pair correlation function (PCF), a surrogate of criticality, was identified, and then *S* was fixed with the value corresponding to the critical point. 20 iterations are conducted for each frequency distribution changing initial phase (r) and amplitude (θ) to find the coupling strength that maximizes the PCF.

#### 2.5.2 Modulation of model FDA

The existing methods to control FDA in previous studies have limitations in significantly altering network structure or frequency distributions [42,49], making them hardly usable for modeling with fixed brain network structures. Therefore, in this study, we propose a novel method to finely control FDA while maintaining network structure and frequency distributions. First, the target FDA is determined for a fixed initial network and frequency distributions. Next, nearby average frequencies for each node and the current FDA and are calculated. If the target FDA is stronger than the current value, a node is randomly chosen, and its frequency ω_*i*_ is exchanged with the node having the order of “# of nodes – the order of the selected node’s frequency” in terms of nearby average frequency. For example, if the selected node has the third-largest frequency, its frequency is exchanged with a node having the third-smallest nearby average frequency. On the other hand, if the target FDA is weaker than the current value, a node is randomly selected, and its frequency is swapped with a node having the order of the same order of nearby average frequency. Using this method, the model FDA is adjusted to have amplitudes of 0, 0.25, and 0.5 and we define this as the “model FDA amplitude”.

#### 2.5.3 Sensitivity test

We conducted simulations for each frequency distribution with different FDA by placing the system at the critical state and applying pulsatile stimulation. By measuring the response to the pulsatile perturbation, we could simulate how the brain network sensitively responds to the external perturbations such as sensory input signals. To measure the sensitivity of the model brain network system, global pulsatile perturbations were applied, and the corresponding responses were measured (Fig.1C) [29]. The pulsatile perturbation *u*(*t*) was added to the amplitude equation, as shown in the following equation.

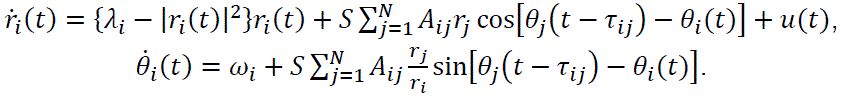

An iteration lasted 25 sec to exclude transient effects (10 sec) and get a sufficient time for baseline estimate (10 sec) with about 100 alpha wave cycles. The pulsatile perturbation was given at 20 sec and lasted 100 ms to include a complete cycle of alpha oscillation. The amplitude of the pulsatile perturbation was 50 to give a perturbation that results in a measurable response without saturating the system. The simulations were done on 100 different initial frequency configurations and 20 iterations with different initial phase and amplitude. A perturbation response time series of ith node PR_*i*_(*t*) can be generated by applying a significance threshold of the 95th percentile to the amplitudes observed in the 10 seconds preceding the onset of stimulation. The values of PR_*i*_(*t*) are set to 1 for amplitudes exceeding the threshold and 0 otherwise. In other words, if the amplitude after stimulation exceeds the 95th percentile of the baseline, we consider it as a responded activity to the stimulus. Responsivity and complexity are defined as 〈PR_*i*_(*t*)〉 and Lempel-Ziv complexity [33] of PR_*i*_(*t*) respectively. Responsivity measures how much quantity responded, and complexity measures how complex the patterns of the responses are. Responsivity and complexity are calculated using 300 ms of PR_*i*_(*t*) from the moment the pulsatile stimulation ends. The large responsivity and complexity after the stimulation means large sensitivity.

#### 2.5.4 Simulation on the frequency of close crisis state

We performed modeling on the occurrence frequency of model VOCs to investigate the relationship between the ES strength and the frequency of VOC occurrence (Fig.1D). Each simulation step lasted for 20 seconds, with normally distributed random constant perturbations (σ=5% of coupling strength at a critical point) given to the network coupling strength *S* in each period. That is, *S* in the Stuart-Landau equation is replaced by *S* × [1 + N(0, 0.05^2^)]. The FDA of each step was calculated using the median frequency of the last 10 second interval and DTI based anatomical network. This is referred to as the “signal FDA amplitude”. Based on the results of EEG analysis, a state with signal FDA amplitude greater than 0.2 was defined as a “close crisis” state. We then counted the occurrences of the close crisis state among 100 steps. We conducted simulations of 20 brain networks with different initial frequency configurations in this scheme.

### 2.6 Statistical analyses

Statistical analysis was performed using either SPSS 29.0.1 or MATLAB. Demographic data were displayed as median and interquartile range. Correlation analyses between frequency disassortativity and each of the PROMs were performed including age and sex as control covariables. Linear regression was constructed including frequency disassortativity as a dependent variable, “close to or distant from VOCs,” and age and sex as predictors. P-values less than 0.05 were considered significant.

## 3. Results

### 3.1 Patient Demographics

Thirty-two participants with SCD with eighteen age-, gender– and ethnicity-matched HCs underwent EEG recording following screening visit and enrolled on the study. PROMs were collected followed by EEG recordings within 1 week. Assessment of recording quality further excluded recordings from 7 participants (Supplementary Figure 1). Study criteria for SCD was listed in Supplementary Table 1. Demographic information was displayed in Table 1. Subjects with SCD and HCs did not differ in age, gender, height, and weight. Participants with SCD showed significantly higher levels of WBC, RBC, Hgb, Hct (%), Reticulocytes (%) and Hemoglobin A (%). Three participants were on chronic transfusion. Fifteen participants were receiving hydroxyurea. In line with the literature, subjects with SCD showed elevated pain, pain interference, depression and physical dysfunction as compared to HCs (Table 1).

**Table 1.** Demographics.

| **Subject Characteristics** | **SCD (n=25)** | **Healthy (n=18)** | **P-value** |
| --- | --- | --- | --- |
| Age (years), median (range) | 37.00 (27.00-45.00) | 38.00 (25.75-57.00) | 0.4562 |
| Female, n (%) | 15 (60.00) | 8 (44.44) | 0.3246 |
| Height (cm), median (range) | 170.1 (164.4-176.5) | 172.6 (167.1-180.8) | 0.3883 |
| Weight (kg), median (range) | 72.40 (59.60-88.40) | 78.30 (67.83-87.55) | 0.2730 |
| **SCD Type Diagnosis** |  |  |  |
| SS/SC/SB0/SB+ (n/n/n/n) | 12/8/4/1 | -- |  |
| **Hematological Indexes** |  |  |  |
| WBC (k/cumm), mean ± SD | 9.23 ± 3.12 | 5.12 ± 1.55 | **<0.0001\*\*\*\*** |
| RBC (million/cumm) , mean ± SD | 3.13 ± 0.78 | 4.67 ± 0.61 | **<0.0001\*\*\*\*** |
| Hgb (GM/dL), mean ± SD | 9.96 ± 1.94 | 13.44 ± 1.54 | **<0.0001\*\*\*\*** |
| Hct (%), mean ± SD | 28.88 ± 5.63 | 40.02 ± 4.30 | **<0.0001\*\*\*\*** |
| Reticulocyte Count (%), mean ± SD | 5.76 ± 3.67 | 1.17 ± 0.43 | **<0.0001\*\*\*\*** |
| Hemoglobin A %, mean ± SD | 24.07 ± 13.02 | 94.79 ± 9.68 | **<0.0001\*\*\*\*** |
| Hemoglobin S %, mean ± SD | 64.71 ± 18.05 | -- | |
| Hemoglobin F %, mean ± SD | 9.46 ± 7.89 | -- | |
| **Disease-modifying Therapy** |  |  |  |
| Chronic transfusion, n (%) | 3 (12.00) | -- |  |
| Hydroxyurea, n (%) | 15 (60.00) | -- |  |
| **Patient-Reported Outcome Measures** |  |  |  |
| BPI Pain Severity | 3.854 ± 2.00 | 0.097 ± 0.4125 | **<0.0001\*\*\*\*** |
| BPI Pain Interference | 3.85 ± 2.12 | -- | |
| FPS Widespread Pain Index | 5.28 ± 3.02 | 0.78 ± 1.59 | **<0.0001\*\*\*\*** |
| HADS Depression Score | 4.76 ± 3.09 | 1.94 ± 2.01 | **<0.0021\*\*** |
| PROMIS-29 Physical Function | 8.40 ± 3.82 | 4.50 ± 1.89 | **<0.0001\*\*\*\*** |
| PedsQL Total Score | 54.36 ± 11.61 | -- | |

### 3.2 SCD subjects have lower median alpha frequencies in the resting state with eyes-closed

We first analyzed EEG data collected from 25 individuals with SCD and 18 HCs during an eyes-closed resting period. The average spectral power between the two groups showed that the alpha peak in SCD is shifted towards lower values (Fig.2A). The shift in median alpha frequency occurred across the entire brain, and it was not localized to any specific regions. The SCD group exhibited a significantly lower (p<10^-5^) frequency of alpha waves (9.01±0.09Hz, Fig.2B) compared to the control group (9.83±0.13Hz, Fig.2B).

**Figure 2.**
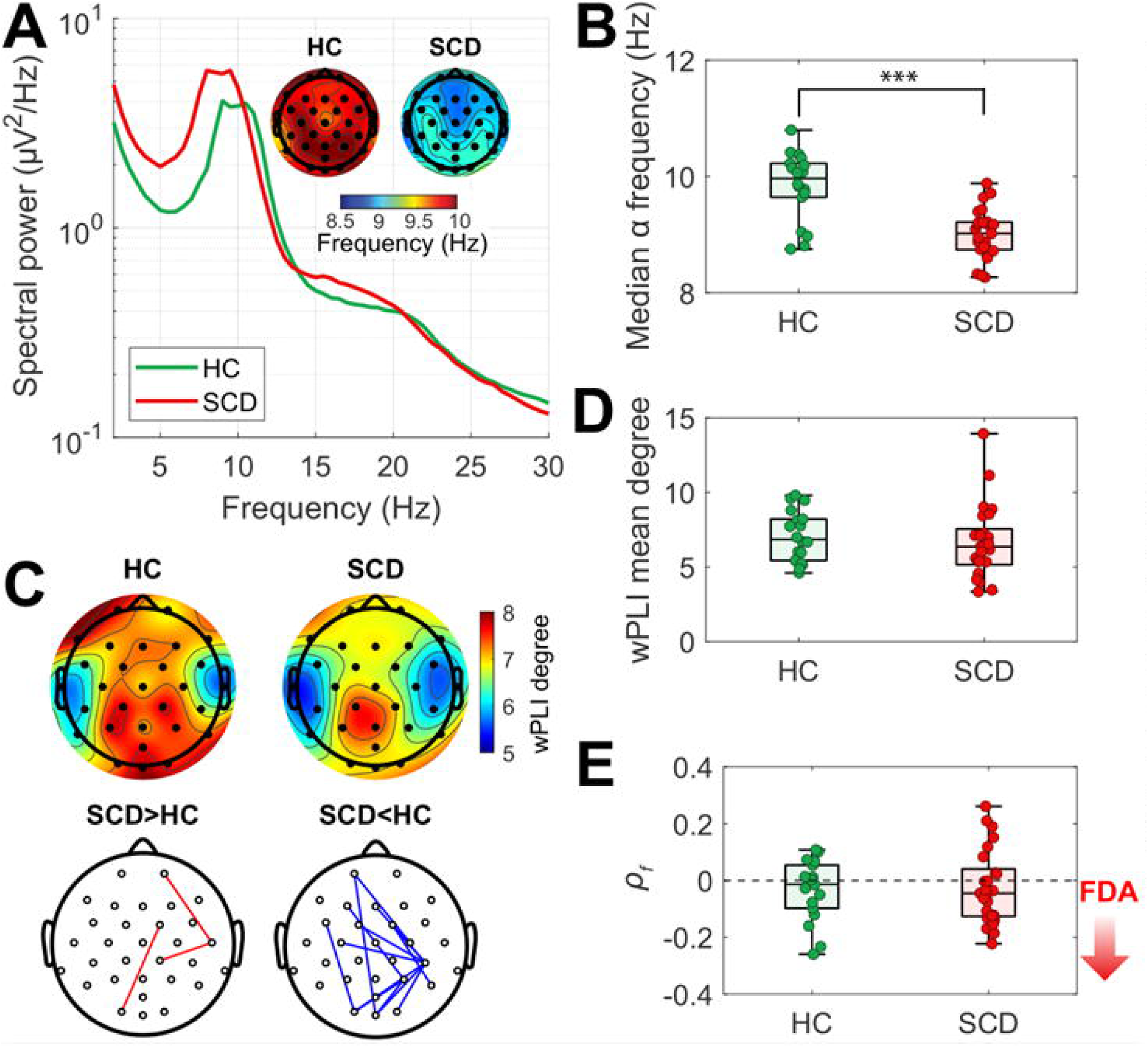
EEG characteristics in eye closed resting state. A) EEG spectral power and topographic plots showing median alpha (7-13 Hz) frequency. B) Comparison of average median alpha frequencies between the HC and SCD groups. SCD group showed significantly lower median alpha frequency (p<10^-5^). C) Topographic plot displaying wPLI degree and the significant difference of wPLI between HC and SCD. wPLI degree is calculated as the sum of weights attributed to the node. There was no significant difference in wPLI degree. Significantly stronger or weaker pairwise wPLI in SCD was indicated by red and blue lines, respectively. D) Mean values of node-wise wPLI degree was not significantly different. E) FDA was not significantly different. ρ_*f*_ is Spearman correlation between median alpha frequencies and the average median alpha frequency of the connected nodes in the binarized wPLI network. The boxplots include horizontal lines indicating the 100%, 75%, 50%, 25%, and 0% percentiles. The statistics were derived from a two-sample t-test.

We calculated wPLI degree as the sum of weights attributed to the node, and there were no significant differences in the node-wise degree and mean degree of wPLI (Fig.2C, D). However, in the pairwise wPLI, 3 pairs were significantly stronger, and 18 pairs were significantly weaker in SCD compared to HCs (Fig.2C). To investigate if there is a difference in FDA between the two groups, we calculated the Spearman correlation ρ_*f*_, and there were no significant differences between the SCD and control groups (Fig.2E). Also, we could observe similar results under pain stimulation. The median alpha frequency significantly differed between HC and SCD groups (p<10^-4^) and SCD showed significantly lower wPLI for 14 pairs (Supplementary Fig. 2).

### 3.3 PROMs of SCD patients are associated with FDA during evoked pressure pain stimulation

The correlation between PROMs and FDA was examined for both resting and during cuff pain, using a partial correlation model that adjusted for age and sex. Partial correlations were made between frequency disassortativity and each PROMs: pain, depression, physical function, and pain-related quality of life. No significant correlations were found during resting state (all p>0.05). In contrast significant correlations were observed during evoked cuff pain, where we found that ρ_*f*_ displayed a significant negative Spearman correlation with: BPI Pain Interference score (ρ=-0.424, p=0.039), PROMISE29 Physical Function (ρ=-0.510, p=0.011), and HADS Depression (ρ=-0.436, p=0.033) in the SCD group (Table 2). The negative correlation between PROMs and ρ_*f*_ suggests that patients with greater pain, depression and poor physical function displayed greater ES features (Stronger FDA).

**Table 2.** Correlation between frequency disassortativity and patient reported clinical outcomes.

| Patient reported outcomes | Spearman correlation (p-value) |
| --- | --- |
| BPI Pain Interference Score | -0.451 (0.031) * |
| BPI Pain Severity Score | -0.336 (0.117) |
| FPS Widespread Pain Index | -0.299 (0.166) |
| HADS Depression Score | -0.482 (0.020) * |
| PROMISE 29 Physical Function Score | -0.620 (0.002) ** |
| PedsQL Total Score | 0.364 (0.088) |

Note: Partial correlation model was constructed for frequency disassortativity and each of the patient-reported outcomes in pain, depression, physical function, and pain-related quality of life, with including age and sex as covariables.

### 3.4 FDA during cuff pain is related to the occurrence of VOCs

We investigated the relationship between ES strength and VOCs by correlating the strength of FDA during cuff pain with the temporal proximity of VOCs. We classified the data into two groups based on time difference between the occurrence of crises and the EEG recording. If one or more crises occurred within 30 days before or after the EEG measurement date, that participant was assigned to the “close crisis” group, and if not, they were assigned to the “distant from crisis” group. We found that a statistically significant Spearman correlation coefficient between ρ_*f*_ and number of VOCs in the previous 12 months (ρ =-0.592; p=0.002). Patients with a higher frequency of VOCs in the previous 12 months displayed stronger FDA (Fig.3A). Importantly, the timing of VOC occurrence relative to EEG recordings was also significantly associated to the ES strength. The group of SCD patients that were in the “close crisis” group showed significantly stronger FDA compared to the “distant from crisis” group (Fig.3B, Table 3). This implies that the closer the VOC occurrence is to the EEG recording, the stronger FDA.

**Figure 3.**
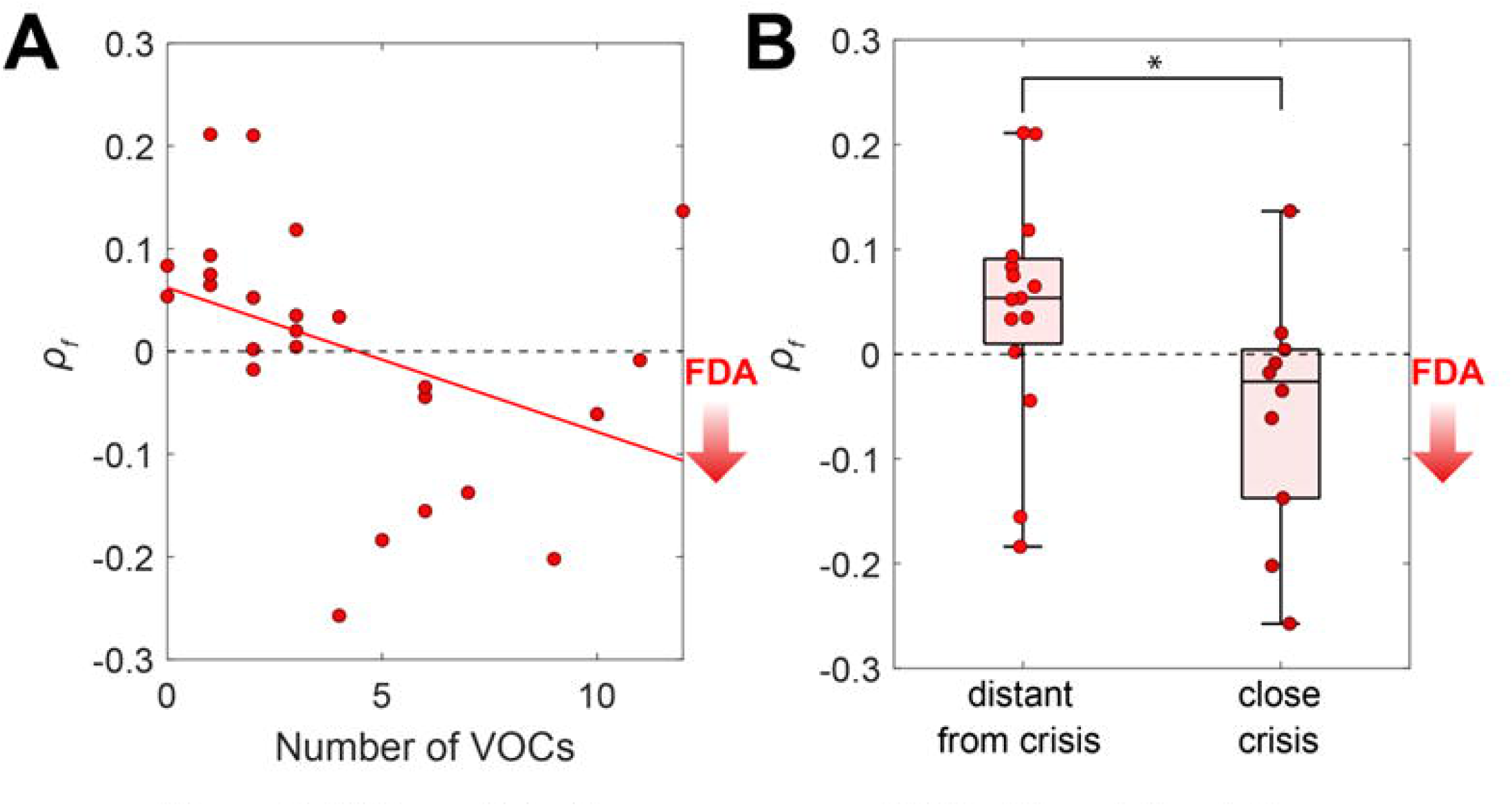
FDA predicts the occurrence of VOC. The relation between FDA calculated from EEG under stimulation and the occurrence of VOCs was analyzed. A) The correlation between FDA and the number of VOCs in the previous 12 months was examined. A significant correlation was found between the number of VOCs and ρ_*f*_. The red line indicates a linear regression. B) The “close crisis” group showed significantly stronger FDA (p=0.035) compared to the “distant from crisis” group. The boxplots include horizontal lines indicating the 100%, 75%, 50%, 25%, and 0% percentiles.

**Table 3.**
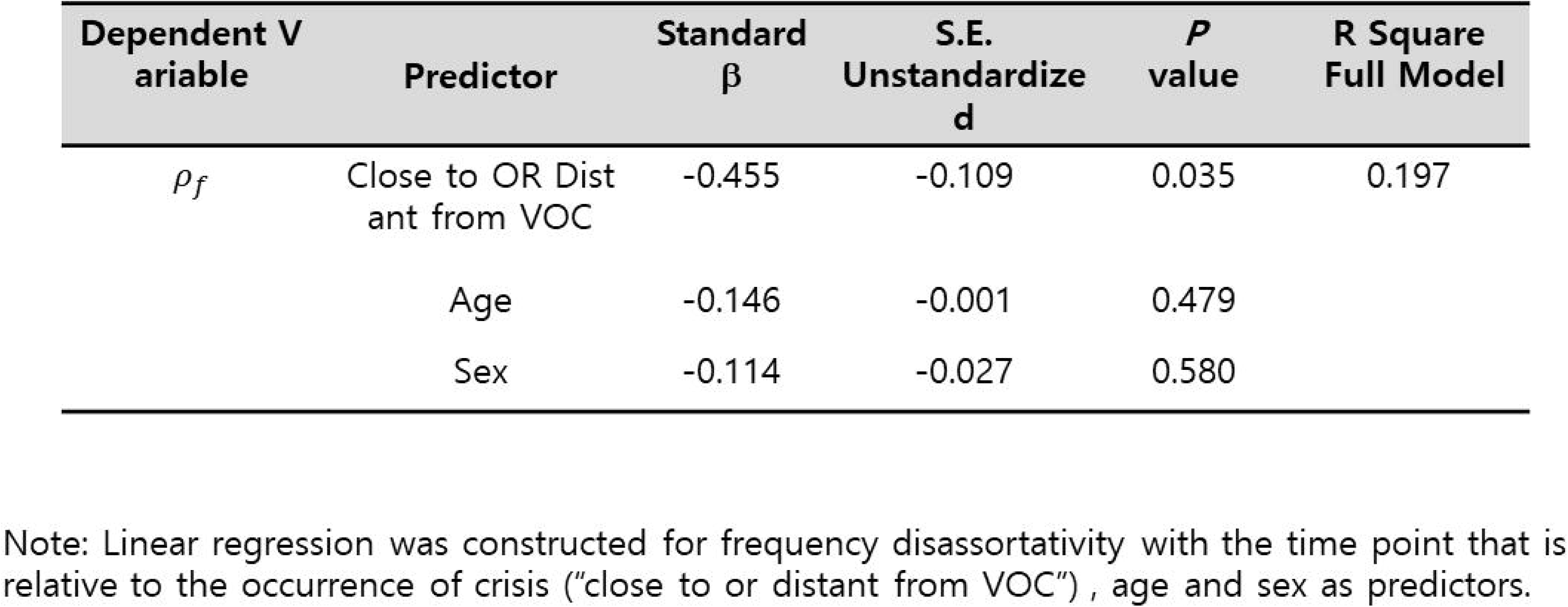
Frequency disassortativitypredicts the occurrence of voe.

### 3.5 Stronger ES strength leads to a more sensitive brain network and increased frequency of painful crises

To investigate the relationship between FDA and network sensitivity, we performed computational modeling on brain network dynamics. Using the DTI-based anatomical brain network consisting of Stuart-Landau oscillators, the responsivity and complexity under pulsatile stimulation were analyzed while varying FDA. Stronger model FDA was associated with higher responsivity and complexity of the model brain network after external perturbation (Fig.4), indicating that the model brain network exhibited increased sensitivity as it approached the strong ES.

**Figure 4.**
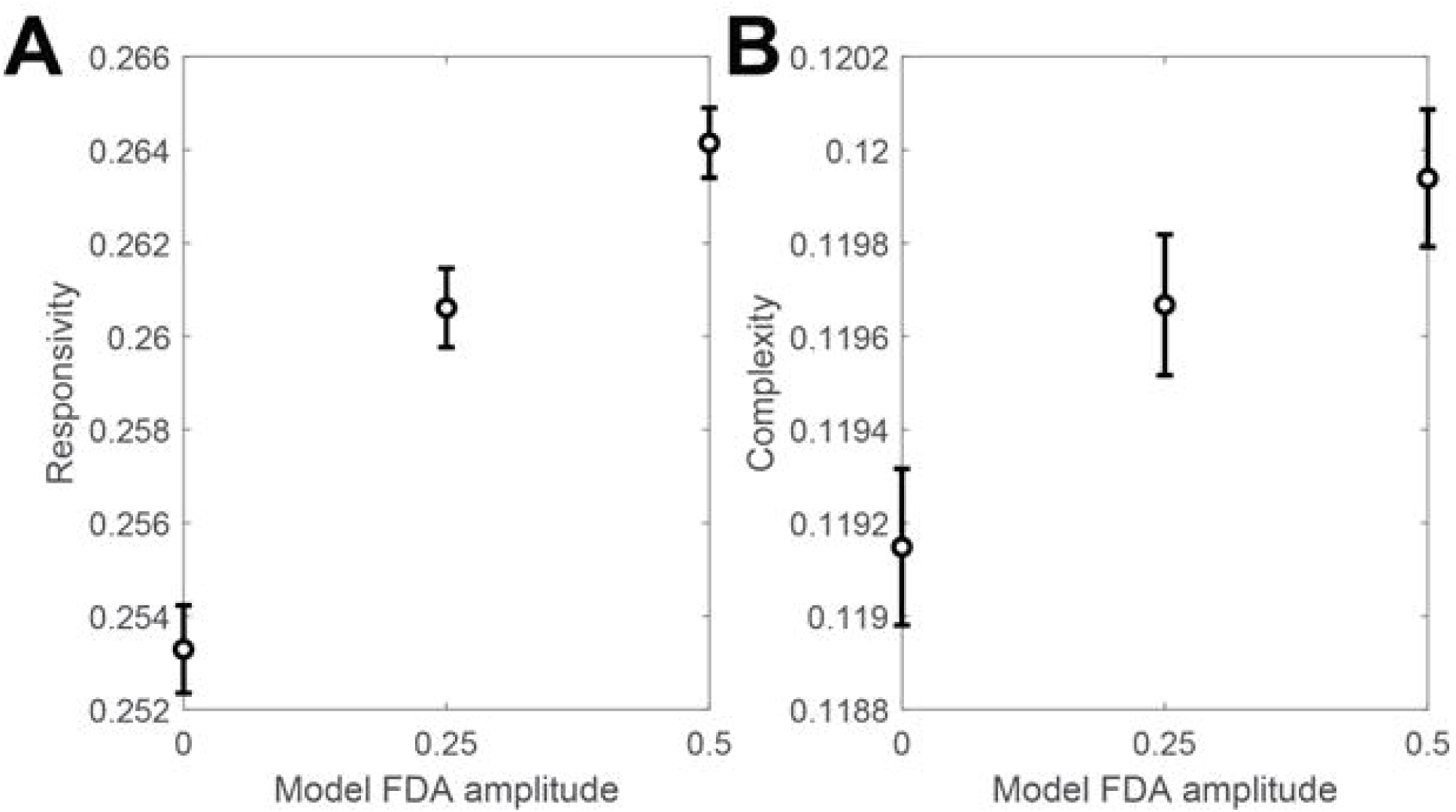
Relation between FDA and network sensitivity. A simulation was conducted to test the sensitivity of the model brain network. Global pulsatile stimulation was applied to the model brain network, and responsivity and complexity were measured from the perturbation response time series PR_*i*_(*t*). A) The responsivity of the brain network to global pulsatile stimulation increases as FDA strengthened. B) The Lempel-Ziv complexity of response time series after global pulsatile stimulation increases as FDA strengthened. The Error bar represents the standard error of the mean.

To examine the relationship between the occurrence of VOCs and FDA, we simulated how brain networks with weak and strong ES strengths fluctuate in response to random perturbations on network coupling strength. Each model network received different perturbations at each 20-second step, and we observed changes in FDA amplitude over time (Fig.5A). We observed that the frequencies of model “close crisis” states were higher when the initial FDA before perturbation was stronger (Fig.5B), indicating that stronger FDA in the model was associated with a higher frequency of VOC occurrence.

**Figure 5.**
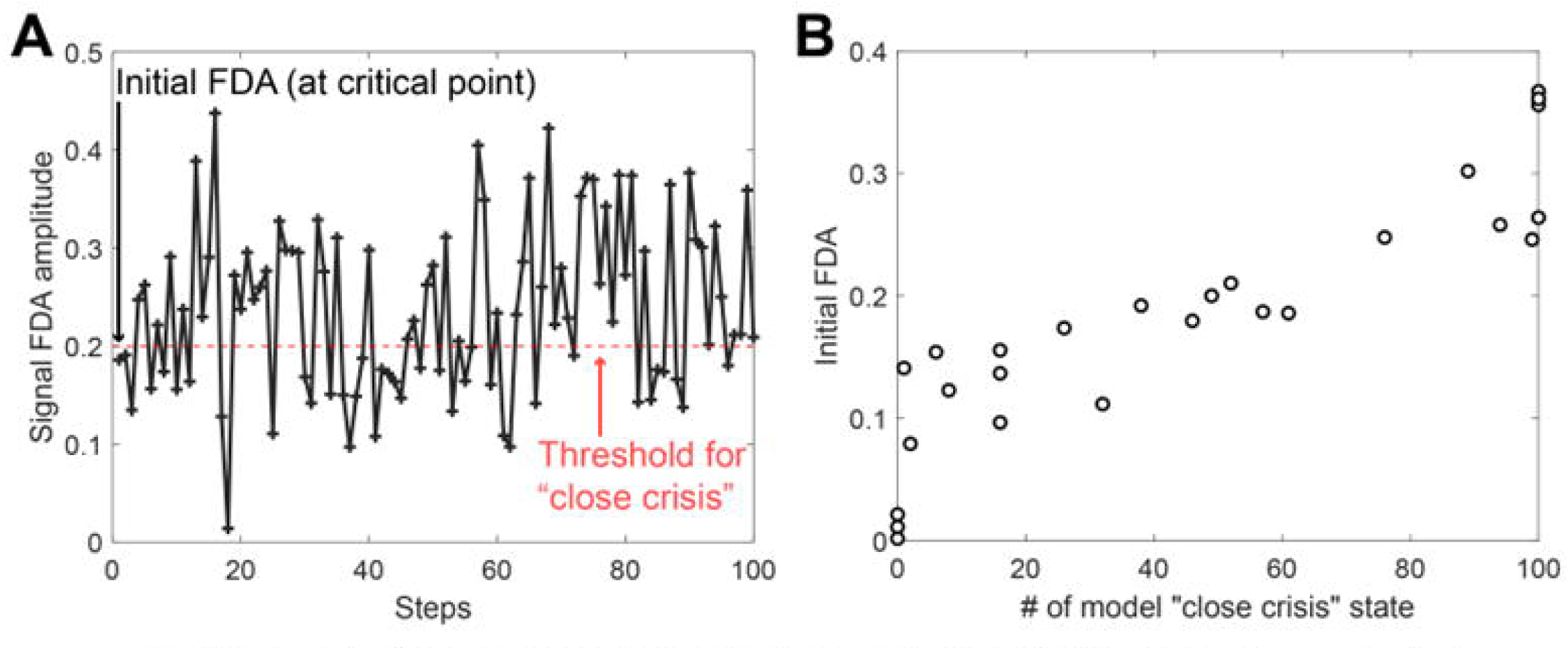
FDA of simulated signal under random perturbation. We conducted simulations to observe how random fluctuations in the coupling strength of the brain network can lead to transitions to states of strong FDA. A) An example of stepwise simulation. Random perturbations are added to the coupling strength in the brain network for each step. B) A higher initial FDA results in a more frequent incidence of model “close crisis” states.

## 4. Discussion

### 4.1 Overview of Results

We analyzed EEG data from individuals with SCD with chronic pain and pain-free age– and gender-matched HCs at rest and during evoked experimental pressure pain. As seen in other chronic pain cohorts [16,17,46], the SCD group exhibited a lower frequency of alpha waves compared to controls. No significant differences were found in the node-wise wPLI degree, but the pairwise wPLI exhibited lower values mainly in SCD. There were no significant differences in FDA between the groups during eyes-closed resting. However, during evoked pressure pain, the FDA of alpha waves in SCD were correlated with levels of pain interference, physical function, and depression. Patients with higher VOC frequency in the past 12 months also presented with stronger FDA, and closer occurrence of crises resulted in stronger frequency disassortativity in the SCD brain network.

We also sought to model the SCD brain network to examine FDA and its relationship to network instability. Computational modeling suggested that stronger FDA in our model was associated with higher responsivity and complexity, which indicated the increased sensitivity to the strong ES strength. We also investigated the occurrence frequency of VOCs by generating random fluctuations to the brain network. Interestingly, stronger initial FDA in the brain network led to higher VOC occurrence in our model, suggesting that the stronger frequency disassortativity may be associated with increased VOC occurrence in individuals with SCD.

### 4.2 The mechanisms of VOC and chronic pain in SCD at a brain network level

The criticality hypothesis suggests that the brain in a conscious resting state resides near a critical state, which is a balanced state between order and disorder or between integration and segregation [1,44]. This means that the brain in a resting state is neither ordered (integrated) nor chaotic (segregated); therefore, it may be able to process information efficiently and effectively [28,37,41]. In thermodynamic systems, there are two types of phase transitions at critical points: the first-order phase transition (ES in a network) and the second-order phase transition (non-ES in a network). ES networks are characterized by higher sensitivity to stimuli and abrupt state transitions compared to lower sensitivity and gradual state transitions in non-ES networks [2]. In patients with SCD, we found that the degree of ES strength is positively correlated with self-reported pain measures and other related symptoms. We also found that as patients approach the onset of VOCs the degree of ES strength is also increased. Finally with computational modeling we demonstrated that the increased ES strength observed in SCD patients may enhance brain sensitivity as well as the frequency of crisis occurrence.

The accumulated effects of SCD within intervals between VOC occurrences may impact specific neural circuits and/or local brain regions altering the global brain network. Previous fMRI studies of SCD have consistently reported altered connectivity in the DMN, a primary hub structure in the global brain network [9,13,23]. Because the brain hub structure is a highly connected and centralized collection of nodes [22,40], we postulate that they are vulnerable to network attacks of accumulating neurobiological factors which may be operative in the SCD brain. Thus, an SCD patient in-between VOCs may accumulate neurobiological factors that promote ES properties mainly in hub structures (e.g., DMN), progressively developing an ES strength until the next VOC.

One possible explanation for the presence of strong ES in SCD patients could be due to weakened brain network efficiency. In line with our observation in this study, it is known that SCD patients display reduced efficiency and in their brain networks [7], which can lead to suppressed global synchronization and the potential emergence of strong ES within the network [2]. That is, weakened efficiency in brain connectivity may paradoxically contribute to the development of a hypersensitive network and excessive pain in SCD. In this study, we observed that the ES strength close to VOCs was stronger compared to a distant one. This may also suggest that as SCD patients approach VOC, the ES characteristics of their brain network become stronger. Also, our model demonstrates that strengthened frequency disassortativity enhances the sensitivity of the brain network, resulting in greater fluctuations in response to external perturbations. Consequently, this heightened sensitivity and increased fluctuations may raise the probability of encountering hazardous states like VOCs. Therefore, the larger variance in FDA within the SCD group and the frequent occurrence of crises among SCD patients can be attributed to the presence of a brain network that exhibits strong ES. The findings in this study provide support for our hypothesis that the increasing strength of ES in the SCD EEG network is associated with the enlarging pain intensity observed during VOC progression (Fig.6).

**Figure 6.**
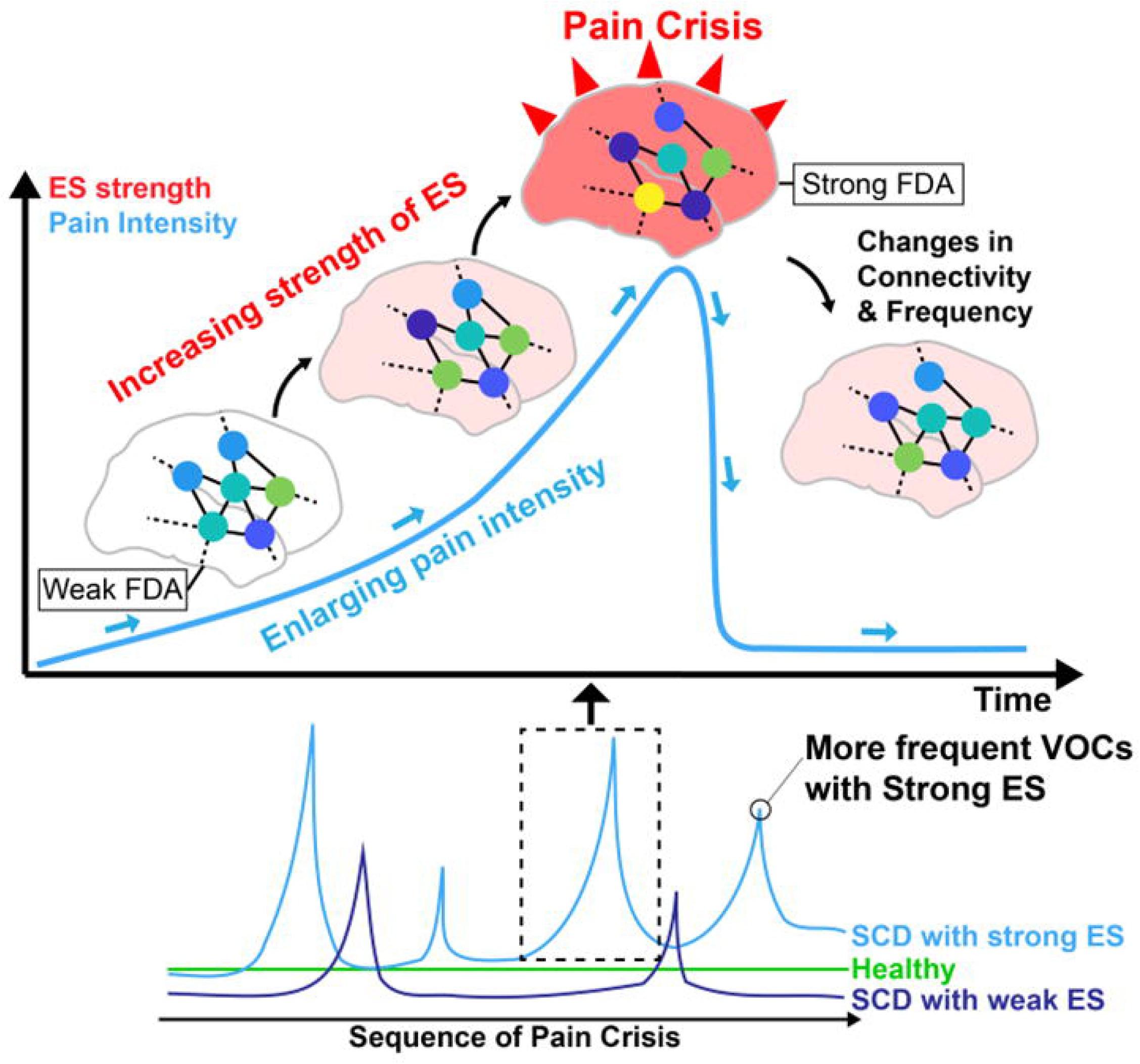
Hypothesis on the network mechanism of pain and VOCs in SCD. The effect of SCD may gradually accumulate within the brain network, strengthening FDA, an ES condition. The cumulative effect on ES can enhance the sensitivity of brain network and upon reaching a critical threshold, potentially triggering a crisis upon reaching a critical threshold. The difference in node colors in the brain network indicates the difference in node frequencies.

We found significant relationships only during the stimulation period, which could be attributed to variations in criticality. We assumed that the brain network in a conscious state resides at a critical point. However, recent empirical evidence suggests that the brain in a conscious resting state may not always stay at the exact critical point. Instead, it slightly deviates towards the sub-critical state (i.e., chaotic and segregated state) [38]. This prevents the brain from a sudden transition to a highly synchronized state (e.g., a seizure) by stochastic internal/external noises. Brain imaging studies have demonstrated that pain stimulation makes functional connectivity within parts of the brain stronger and integrative [23,39], and generally, increasing functional connectivity in a network can play a role in pushing a system in a sub-critical state (a segregated state) back closer to its critical point. Therefore, the pain stimulation may push the SCD brains closer to their critical points, making the difference between ES and non-ES more pronounced when they are near their critical points.

### 4.3 Explosive synchronization (ES) may be a novel marker of VOC onset and modulation

Over the last decade, many network configurations that can induce ES have been discovered, including: (1) positive correlation between node degrees and node frequencies, (2) negative frequency assortativity (a tendency of high-frequency nodes to be linked with low-frequency nodes), (3) large frequency difference, (4) random connectivity, and (5) negative feedback process among nodes [2,15,25]. Our findings indicate the potential of FDA as a predictor for VOCs in SCD. FDA was first shown to have a correlation with pain in fibromyalgia [32]. Here, we observe positive correlations between the strength of ES and symptoms of pain and depression in SCD patients, while no significant correlations were found for other ES conditions. The consistency of FDA across diverse pain conditions, including fibromyalgia and SCD, implies its potential as a predictive measure for pain, irrespective of underlying mechanisms of pain. This may allow the finding of better ES conditions that can reflect the pain scores and predict the upcoming pain crisis. This needs further study to develop the network principle-based prediction system of pain crises.

### 4.4 ES can provide a theoretical framework for the treatment of SCD pain through network modulation

Discovering correlations between ES strength in the SCD brain, pain intensity, and frequency of VOCs could help us develop a novel treatment method to reduce pain intensity and frequency of pain crises. In a recent computational model, we showed that increasing connectivity in hub regions (e.g., insula, isthmus cingulate cortex, and precuneus) in an ES brain network can convert it into a non-ES brain network [27], which significantly reduces the brain’s sensitivity to external stimuli. Our next step is to apply a stimulation method (e.g., acupuncture or transcranial direct current stimulation) that is known to alter brain connectivity [4,31] to our modeling approach. Based on the computational study and our empirical findings in SCD, we could develop a novel theoretical basis for systematically predicting and modulating pain intensity as well as the frequency of VOCs by converting the type of phase transition in individual SCD patients’ brains.

### 4.5 Limitations

Our study offers valuable insights into brain network mechanisms of VOCs in SCD; however, it also has certain limitations. In analytic and modeling studies, frequency disassortativity in a network is defined by the initial arrangement of frequencies (called natural frequencies) to the nodes. However, we cannot directly measure natural frequencies in brain regions using EEG data. Furthermore, we determined the median frequency from the band EEG, ignoring the spectral distribution. This simplification in sensor signals may not accurately reflect the real-brain network dynamics in source signals. Simultaneous recordings of fMRI and EEG may enable us to estimate the strength of ES more precisely. Finally, the amount of data recorded near the onset of VOCs was also largely restricted by the number of VOCs naturally occurring that were adjacent to the recording time. Due to this limitation of not knowing precisely when a VOC is going to occur prospectively, there is a significant challenge in using EEG to study or clinically manage VOCs.

### 4.6 Conclusion

The present study establishes a significant link between pain in SCD and a universal network mechanism, ES. It offers a robust theoretical foundation for comprehending pain in SCD through the brain network mechanism. This enhanced understanding will facilitate future investigations on predicting pain crises and refining pain management strategies for SCD patients.

## Data Availability

All data produced in the present study are available upon reasonable request to the authors.

## Acknowledgement

The authors would like to thank Andrew Q Pucka, Brandon Alec Reyes, Tyler James Barret, Nayana Dutt, Payton Mittman, and Bea Paras for assisting with experimental performance; Siddhi Gandhi, Charanjit Kaur, and Veena Vijayan for data management and validation, as well as all the providers affiliated with Indiana University Health and Indiana Hemophilia & Thrombosis Center for patient referral to this study. The authors also thank Indiana University Clinical Research Center for resource support.

This work was supported by NIH K99/R00 award (Grant # 4R00AT010012) and Indiana University Health – Indiana University School of Medicine Strategic Research Initiative funding to YW. UL was supported by NIGMS (R21GM143521).

PJ, REH, UL and YW analyzed and interpreted the data and drafted the manuscript; M K assisted with data analyses and interpretation; PJ and UL conducted computational modeling; BK, VV and YT assisted with data collection; SEH, REH, UL, and YW edited the manuscript; YW directed patient recruitment, experimental performance, data collection and quality of the clinical investigation.

---

**Supplementary Figure 1.**
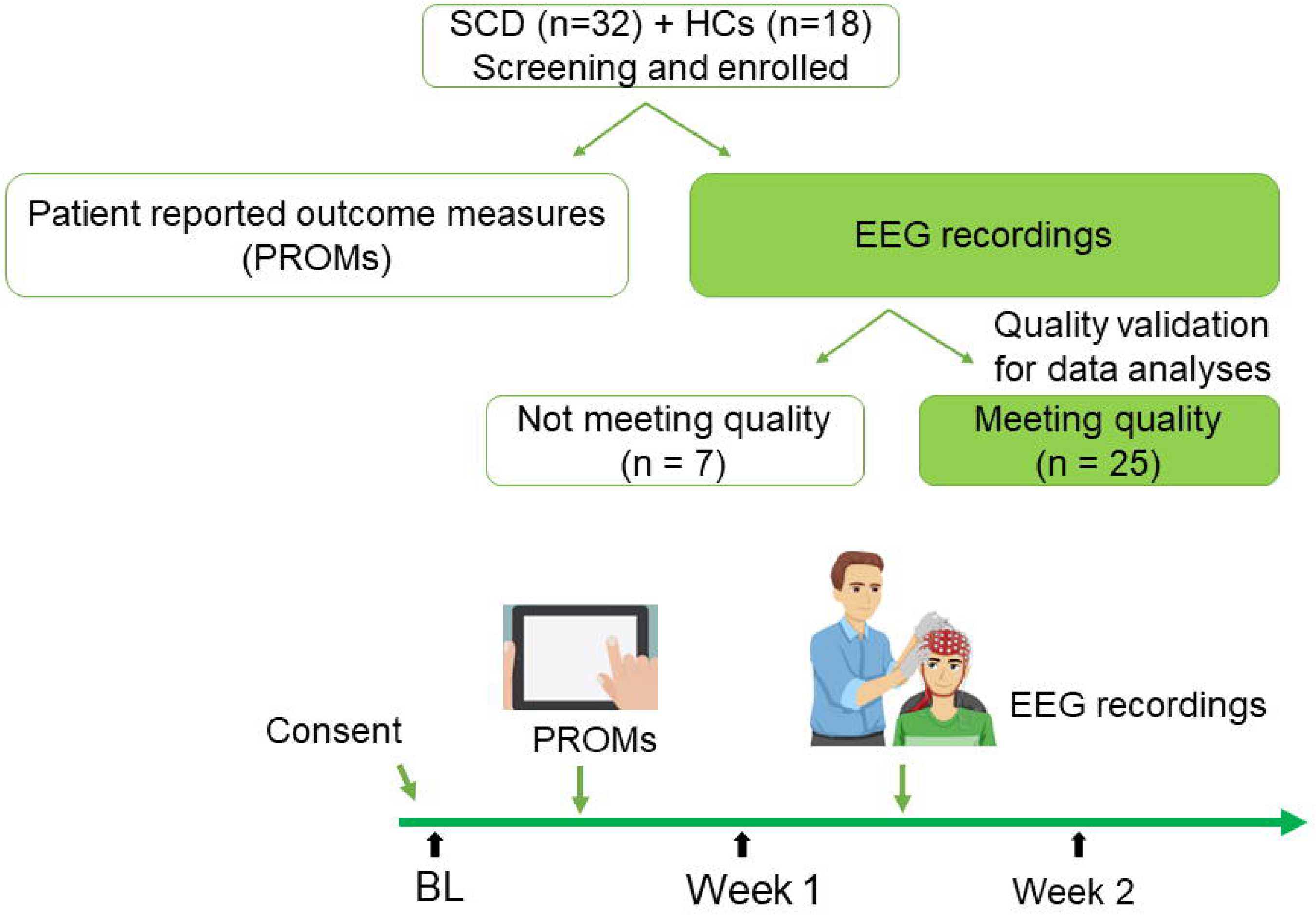
Study workflow of enrollment and analyses.

**Supplementary Figure 2.**
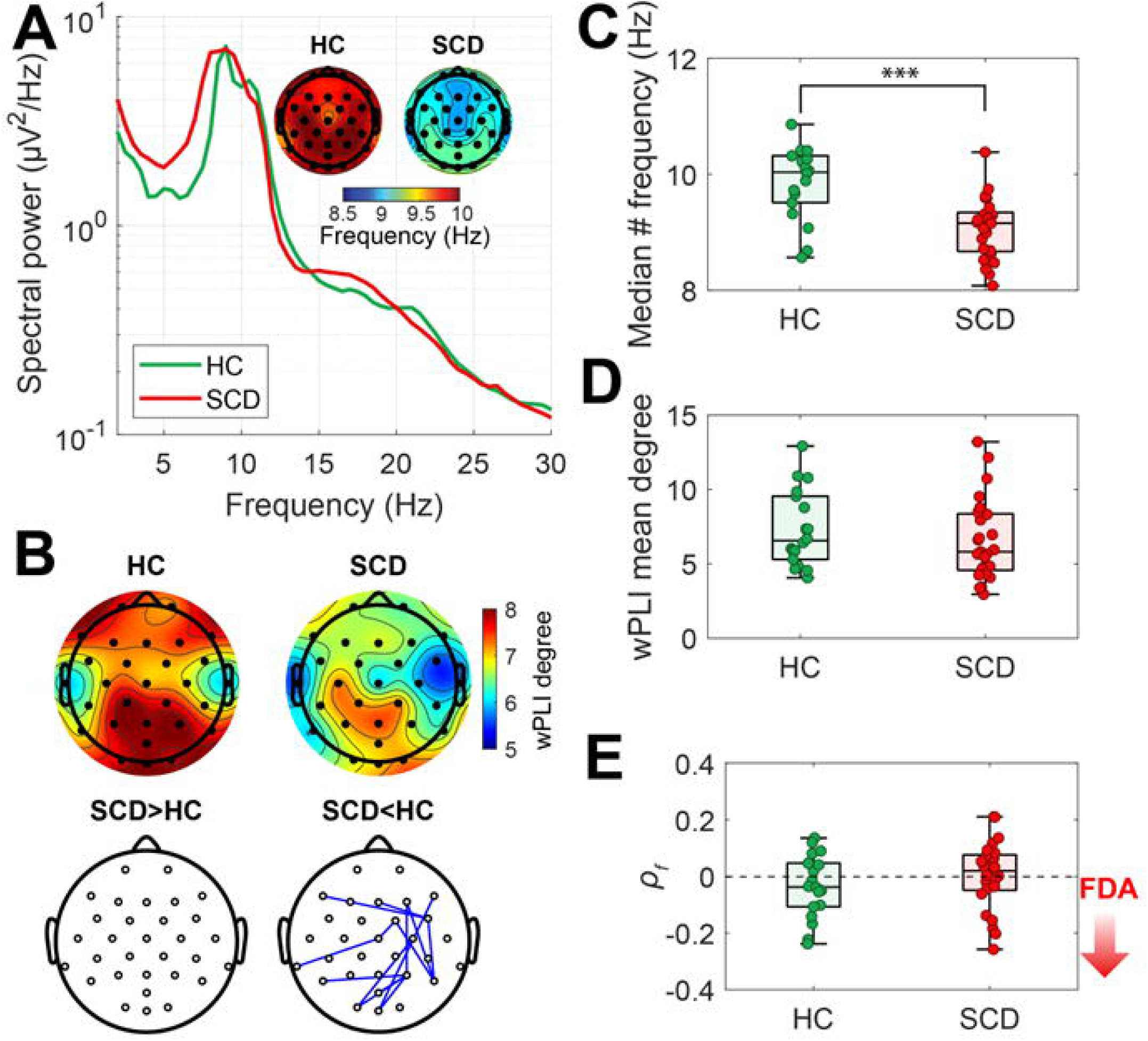
Analysis results of EEG under stimulation. A) EEG spectral power and topographic plots showing median alpha (7-13 Hz) frequency. B) Topographic plot displaying wPLI degree and the significant difference of wPLI between HC and SCD. wPLI degree is calculated as the sum of weights attributed to the node. There was no significant difference in wPLI degree. Significantly stronger or weaker pairwise wPLI in SCD was indicated by red and blue lines, respectively. C) Comparison of average median alpha frequencies between the HC and SCD groups (p<1O·^4^). D) Mean values of node-wise wPLI degree was not significantly different. E) FDA was not significantly different. *P_f_* is Spearman correlation between median alpha frequencies and the average median alpha frequency of the connected nodes in the binarized wPLI network. The boxplots include horizontal lines indicating the 100%, 75%, 50%, 25%, and 0% percentiles. The statistics were derived from a two-sample t-test.

**Supplementary Table 1.**
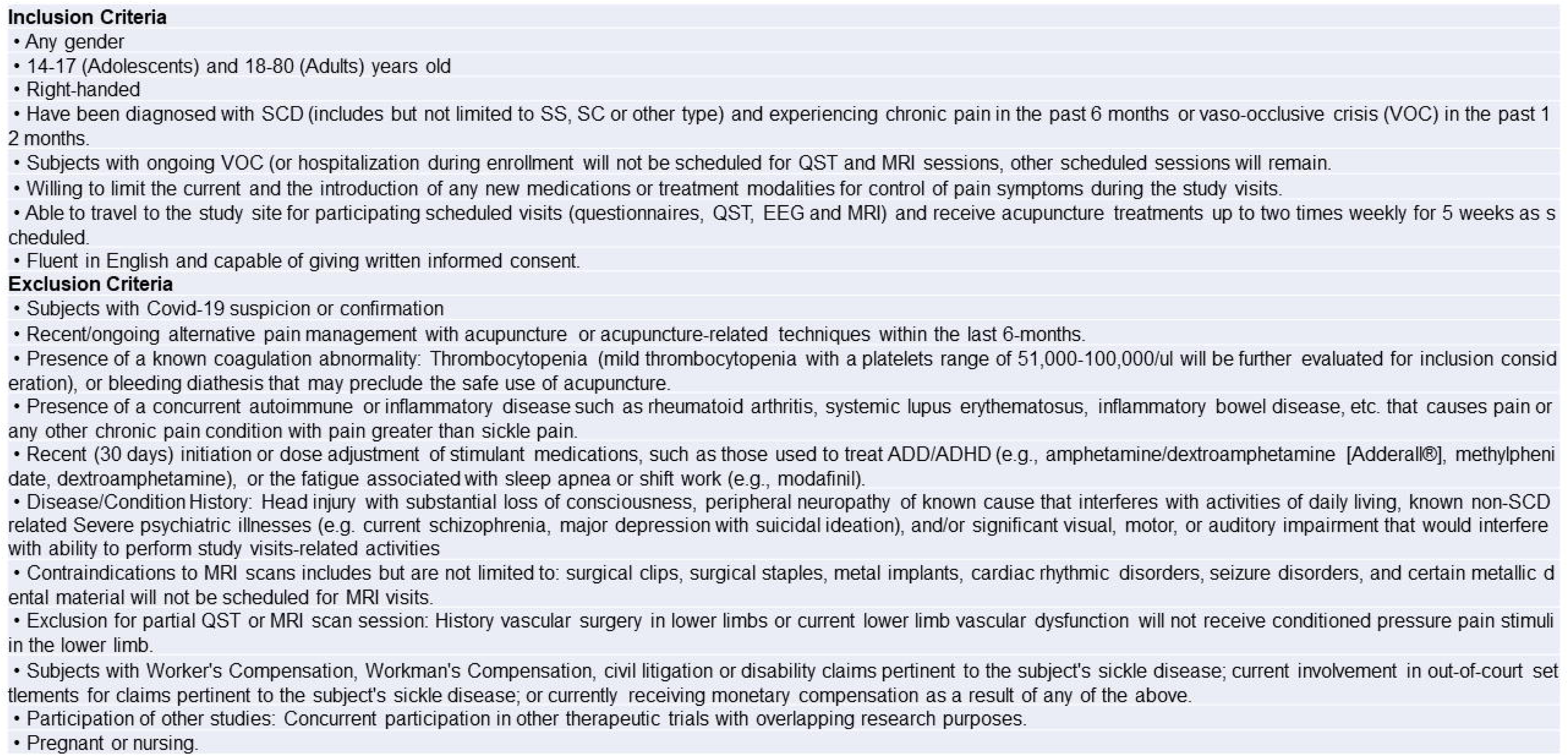
Study Eligibility Criteria.

